# Durability for 12 months of antibody response to a booster dose of monovalent BNT162b2 in adults who had initially received 2 doses of inactivated vaccine

**DOI:** 10.1101/2023.08.18.23294185

**Authors:** Eunice Y. C. Shiu, Samuel M. S. Cheng, Mario Martín-Sánchez, Niki Y. M. Au, Karl C. K. Chan, John K. C. Li, Lison W. C. Fung, Leo L. H. Luk, Sara Chaothai, Tsz Chun Kwan, Dennis K. M. Ip, Gabriel M. Leung, Leo L. M. Poon, J. S. Malik Peiris, Nancy H. L. Leung, Benjamin J. Cowling

## Abstract

We administered BNT162b2 as a third dose to 314 adults ≥30 years of age who had previously received 2 doses of inactivated vaccine. We collected blood samples before the third dose and again after 1, 6 and 12 months, and found stable levels of antibody responses to the ancestral strain and Omicron BA.2 at 6-12 months after receipt of the BNT162b2 third dose, with increased antibody levels in individuals who also received a fourth vaccine dose or reported a SARS-CoV-2 infection during follow-up.

## INTRODUCTION

A number of different SARS-COV-2 vaccine platforms have been widely used since late 2020 to mitigate the morbidity and mortality impact of COVID-19. Inactivated vaccines have been some of the most widely used vaccines, providing effective protection against severe COVID-19 [1,2], but stimulating weaker neutralizing antibody responses after vaccination compared to other vaccine platforms [3,4]. We established a trial to investigate the strength and durability of antibody responses to a heterologous third dose of BNT162b2 (BioNTech/Fosun Pharma/Pfizer) in adults ≥30 years of age who initially received two doses of an inactivated vaccine. We previously reported initial antibody responses to the third dose of BNT162b2 [5] and durability for up to six months [6], and here we report further data up to 12 months after receipt of the third dose and investigate the effect of Omicron infections and fourth doses on antibody levels.

## METHODS

### Study Design

We established a single-arm open label trial in adults who had previously received two doses of inactivated vaccine. Participants were eligible if they were at least 30 years of age, and had received their second inactivated vaccine dose ≥90 days prior to enrollment in this trial.

Participants were ineligible if they had a history of laboratory-confirmed SARS-CoV-2 infection prior to enrolment, if they met a contraindication for receipt of BNT162b2, were receiving immunomodulatory medications, or were females who were pregnant or intending to become pregnant in the upcoming 3 months.

We collected serum samples at enrollment immediately before administration of BNT162b2, and again on day 28, day 182 and day 365. At enrollment we collected information on demographics and self-reported SARS-CoV-2 infection history, and reviewed COVID-19 vaccination records. We updated this information at the day 182 and day 365 visits including information on any infections that had occurred during follow-up or any additional vaccine doses that had been received.

### Ethical approval and trial registration

All participants provided written informed consent to participate. The study was approved by the Institutional Review Board of the University of Hong Kong. The study is registered at ClinicalTrials.gov (NCT05057182).

### Laboratory Methods

Sera was extracted from the clotted blood within 48 hours and stored at -80°C until subsequent serologic testing. We tested serum samples with a surrogate virus neutralization test [7] for which the kits were obtained from GeneScript USA Inc. (Piscataway, New Jersey) and the assays were performed in accordance with the manufacturer’s recommendations. Inhibition levels ≥30% were considered positive. We tested antibody responses to the ancestral strain, Omicron BA.2 and we also tested the day 182 and 365 samples to Omicron BA.5. A random subset of samples were tested with a plaque reduction neutralization test (PRNT) with live ancestral virus and Omicron BA.2 virus [8]. PRNT assays were performed in duplicate using 24-well tissue culture plates (TPP Techno Plastic Products AG, Trasadingen, Switzerland) in a biosafety level 3 facility using Vero E6 TMPRSS_2_ cells. Serial dilutions from 1:10 to 1:320 of each serum sample were incubated with 30-40 plaque-forming units of virus for 1 hour at 37°C. Virus-serum mixtures were subsequently added onto pre-formed Vero E6 cell monolayers and incubated for 1 hour at 37°C in a 5% CO_2_ incubator. The cell monolayer was then overlaid with 1% agarose in cell culture medium and incubated for 3 days, at which time the plates were fixed and stained [7,8]. PNRT_50_ titers were defined as the highest serum dilution that resulted in ≥50% reduction in the number of virus plaques. Positive and negative control sera were included in every experiment. All assays listed above were previously validated and reported [9].

### Statistical Analysis

We presented the sVNT inhibition (%) and PRNT_50_ titers in serum samples collected at day 0, day 28, day 182, day 365 after the administration of the third dose of BNT162b2. Information on SARS-CoV-2 infections and the receipt of any additional booster doses during follow-up were self-reported by participants. We compared the group means for surrogate virus neutralization (sVNT) levels and the geometric mean PRNT_50_ titers in the serum samples between groups using t tests. Statistical analyses were conducted using R version 4.2.2 (R Foundation for Statistical Computing, Vienna, Austria).

## RESULTS

We enrolled and administered BNT162b2 as a third dose to 314 participants between 18 October and 28 December 2021. We successfully followed up 277 (88%) through to the day 365 blood draw, including day 28 samples from 312 (99%) participants and day 182 samples from 284 (90%) participants. The day 365 samples were collected from 26 September 2022 through to 13 February 2023. Participants who completed the day 365 blood collection had a median age at enrolment of 54 years; IQR: 48 -62) and 62% were male. The age and sex distribution of the 12% of enrolled participants who did not provide day 365 blood samples were similar to those who did provide samples at day 365, with a median age at enrolment of 48 years (IQR: 40 -61) and 59% being male.

During the follow-up period, among the 277 participants followed through to day 365, 98 reported receipt of a fourth dose and these were received between April 2022 and December 2022. Among these fourth doses, the majority were with monovalent BNT162b2 (86, 88%), with 11 participants receiving the monovalent CoronaVac and one participant receiving the bivalent formulation of BNT162b2. Two participants reported receiving a fifth dose of bivalent BNT162b2 at 13 and 42 days prior to day 365. 76 participants reported a SARS-CoV-2 infection confirmed by PCR or by rapid antigen test between February 2022 and November 2022, with the majority of these (35, 46%) occurring during the month of March 2022. 20 participants reported both a fourth dose and a confirmed SARS-CoV-2 infection, of which 11 received the fourth dose prior to infection (median days, 33; range, 1-133), and the remaining 9 received a fourth dose after the infection occurred (median days, 176; range, 22-222). One participant received a fourth dose one month after the confirmed infection, and subsequently received a fifth dose 8 months later.

Among individuals without a reported infection or a fourth dose of vaccination, antibody responses against the ancestral and Omicron BA.2 strain were substantially higher in all assays after the third dose, declined by 6 months, and were similar at 12 months with no apparent further decline (Figure 1C and 1D). In comparison, PRNT_50_ titers of the Omicron BA.2 strain were higher at the subsequent blood draw among those who reported a SARS-CoV-2 infection, with the geometric mean PRNT_50_ titer of 128 at 6 months and 69 at 12 months (Figure 1D). In a subset of participants who reported a SARS-CoV-2 infection in addition to the receipt of a fourth dose prior to day 365, the antibody titers against BA.2 were higher at day 365 with the geometric mean of PRNT_50_ titer of 127 as compared to the geometric mean titers of 69 in adults with infection only (p<0.01) and 25 with a fourth dose only (p<0.01) (Figure 1).

**Figure 1.**
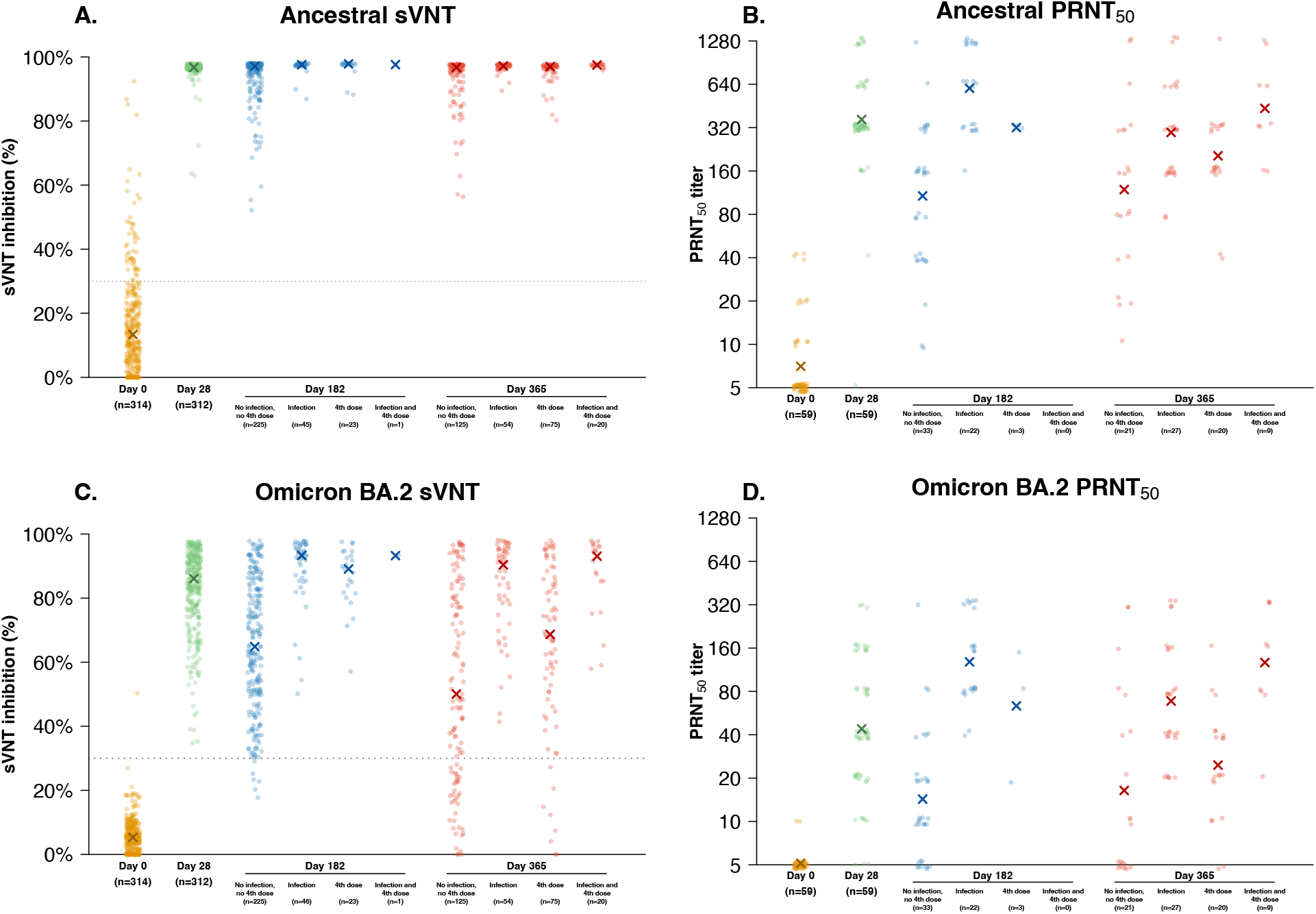
Antibody titers by sVNT and PRNT_50_against live SARS-CoV-2 at day 0, day 28, day 182 and day 365 after receipt of a third dose of BNT162b2 in participants. Analyses at day 182 and 365 are stratified by reported SARS-CoV-2 infection or additional booster dose. Panel A: antibody levels against the ancestral strain by sVNT, with X indicating the median level. Panel B: antibody titers against the ancestral strain by PRNT_50_, with X indicating the median level. Panel C: antibody levels against the Omicron BA.2 strain by sVNT, with X indicating the median level. Panel D: antibody titers against the Omicron BA.2 strain by PRNT_50_, with X indicating the median level. In panels A and C the dotted line at a value of 30% represents the manufacturer’s reported threshold for seropositivity on this assay.

We further examined changes over time in antibody levels against the Omicron BA.2 and BA.5 subvariants in those who reported receipt of a fourth dose of monovalent BNT162b2 or reported an infection. There was a decreasing trend in the PRNT_50_ levels against BA.2 and the sVNT levels against BA.2 and BA.5 in participants from one month up to six months after the receipt of a fourth dose of monovalent BNT162b2 only, or from one month up to six months after infection and without receipt of any additional booster doses (Figure 2). In the group who received a fourth dose, the antibody levels against BA.2 were maintained at high levels after 2 months, but gradually waned to a median PRNT_50_ titer of approximately 20 and a median sVNT level of 55% at 6 months. The antibody levels after reported infection also waned over time, but the median PRNT50 titer remained above 40 and the median sVNT level remained above 60% at 8 months post-infection. There were insufficient participants and data points to examine changes over time in antibody titers after a fourth vaccine dose with CoronaVac or bivalent BNT162b2, or after a fifth vaccine dose.

**Figure 2.**
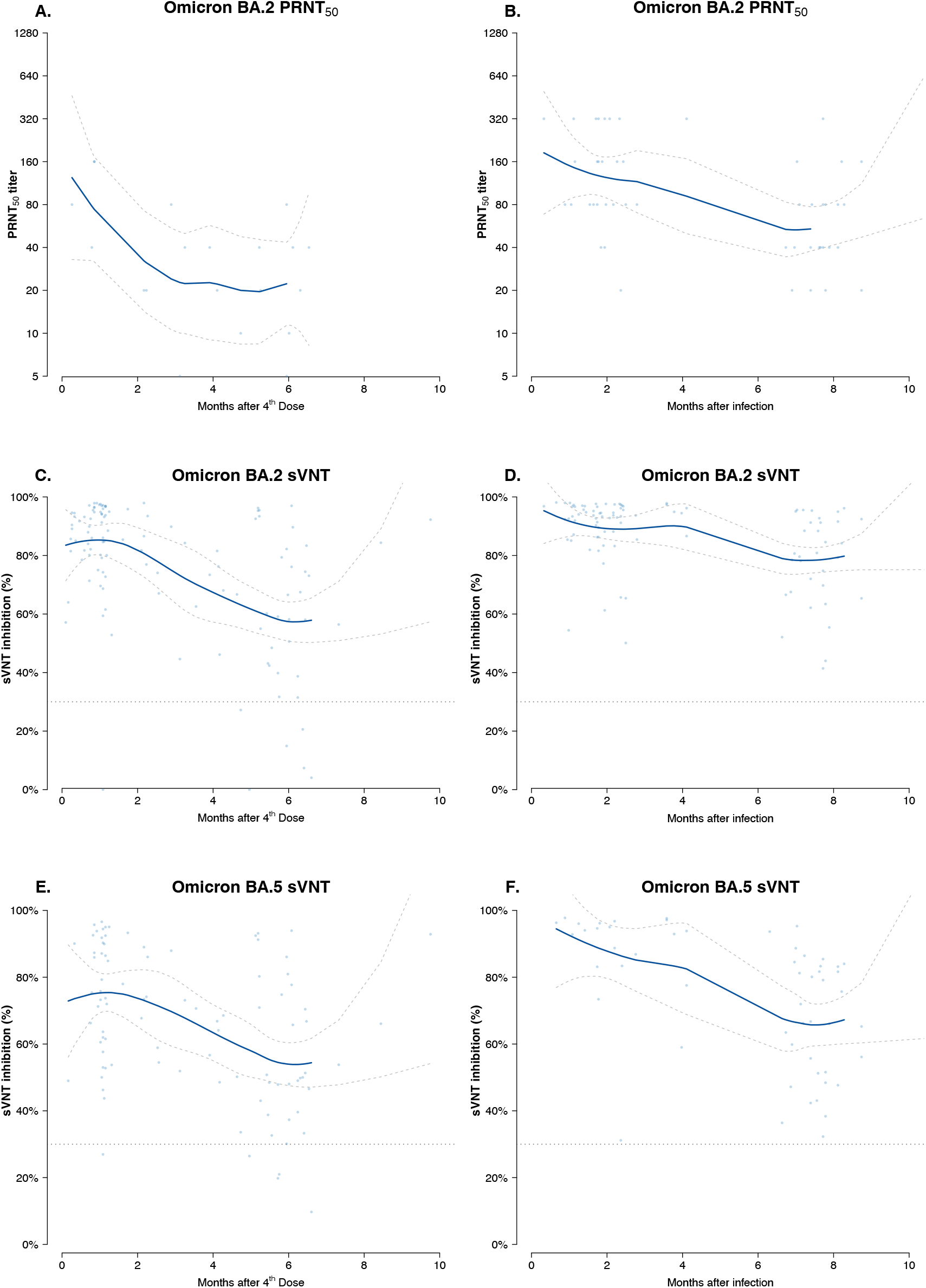
Antibody levels by time since third dose measured by a plaque reduction neutralization test and a surrogate virus neutralization test against Omicron BA.2 and Omicron BA.5 in individuals who reported a SARS-CoV-2 infection or a fourth dose of monovalent BNT162b2 during follow-up. Panel A: PRNT_50_ titers against Omicron BA.2 by time since receipt of a fourth dose of monovalent BNT162b2. Panel B: PRNT_50_ titers against Omicron BA.2 by time since self-reported SARS-CoV-2 infection. Panel C: sVNT antibody levels against Omicron BA.2 by time since receipt of a fourth dose of monovalent BNT162b2. Panel D: sVNT antibody levels against Omicron BA.2 by time since self-reported SARS-CoV-2 infection. Panel E: sVNT antibody levels against Omicron BA.5 by time since receipt of a fourth dose of monovalent BNT162b2. Panel F: sVNT antibody levels against Omicron BA.5 by time since self-reported SARS-CoV-2 infection. In each panels the smooth curve represents the smoothed average and the 95% pointwise confidence intervals are shown as dashed lines to either side. In panel C, D, E and F the dotted horizontal lines at a value of 30% represent the manufacturer’s reported threshold for seropositivity on this assay.

## DISCUSSION

The result of this study show that a third vaccine dose with monovalent BNT162b2 increased antibody levels against the ancestral strain and Omicron BA.2 strain in adults who had received 2 doses of inactivated vaccines, and antibody levels remained above a predicted protective threshold against infection [7] for at least 12 months after the third dose. Antibody levels gradually declined over time since the third dose, but additional booster doses or infections were associated with higher antibody levels at day 365 (Figure 1).

Among participants who received a fourth dose, 88% received monovalent BNT162b2. The monovalent BNT162b2, containing mRNA of the ancestral strain, may provide some degree of protection against the Omicron strains [10]. In our study, the antibody titer against the Omicron BA.2 strain in participants with receipt of a fourth dose of monovalent BNT162b2 was higher than the predicted protective threshold at 6 months and 12 months, but it was comparatively lower than the titers in participants with reported infection, even with a longer delay from infection to day 365 blood collection (median delay from infection, 210 days; median delay from fourth dose, 156). Most infections in our cohort were reported during a large community epidemic of the Omicron BA.2 subvariant in Hong Kong [11], likely explaining why the antibody levels against Omicron BA.2 were much higher in infected individuals than the individuals with a fourth dose of monovalent BNT162b2 (Figure 2).

A relatively small number of participants reported receipt of bivalent BNT162b2 prior to day 365, therefore we were unable to assess the antibody responses from bivalent BNT162b2 here. Our study has several other limitations. We observed an infection rate of 24% in our cohort, which was slightly less than the estimated 41% infection rate during the Omicron wave in Hong Kong [12], although receipt of the third dose in our study between October and December 2021 might have provided some degree of protection against confirmed SARS-CoV-2 infection in March 2022 [10,13]. Only one participant reported two episodes of COVID-19 infection during the 1-year follow up period, but some unrecognized infections/re-infections might have occurred. Finally, while we measured antibody levels, we did not directly correlate these levels with the degree of protection against infection, and this would be an important area for further research.

In conclusion, our study demonstrates that a BNT162b2 booster dose provides durable antibody response in individuals who initially received two doses of an inactivated SARS-CoV-2 vaccine, with participants maintaining high levels of sVNT antibodies at 12 months after receipt of the third dose.

## Data Availability

All data produced in the present study are available upon reasonable request to the authors

## ACKNOWLEDGMENTS

We gratefully acknowledge colleagues including Zacary Chai, Sara Chaotahi, Kelvin Kwan, Yvonne Ng, Teresa So, and Eileen Yu for technical support in preparing and conducting this study; Anson Ho for setting up the database; Julie Au and Lilly Wang for administrative support; Hetti Cheung, Victoria Wong, and Bobo Yeung at the University of Hong Kong (HKU) Health System; Cindy Man and other colleagues at the HKU community Vaccination Centres at Gleneagles Hospital; and all the study participants for facilitating the study.

## DISCLAIMER

The funding bodies had no role in the design of the study, the collection, analysis, and interpretation of data, or writing of the manuscript.

